# A reliability-screened thalamocortical control-network phenotype tracks cocaine-use history in cocaine use disorder

**DOI:** 10.64898/2026.04.28.26351962

**Authors:** Brice Edelman, Jeffrey Skolnick

## Abstract

**Background:** A central goal in psychiatry is to move from symptom-defined diagnoses toward biologically interpretable and reliable phenotypes. In cocaine use disorder (CUD), many resting-state abnormalities have been reported, but few circuit-level findings have been explicitly screened for reliability. We tested whether prespecified thalamocortical features yield a reproducible phenotype in CUD and whether that phenotype reflects diagnosis, recent cocaine use, or longer-term illness history.

**Methods:** Discovery analyses used resting-state data from 105 participants (46 healthy controls, 59 CUD). From a 13-region thalamocortical circuit, we derived an HC-trained LEiDA state model, generated 11 prespecified features, and advanced only those meeting split-half reliability criteria (ICC[3,1] ≥0.40). A separate paired TMS sample (n=44) was used for extension analyses.

**Results:** Five features survived reliability screening. Within CUD, longer duration since beginning cocaine use was associated with greater occupancy of a control-like state (standardized β=0.37, q=0.005) and stronger whole-thalamus connectivity with control frontoparietal cortex (standardized β=0.30, q=0.018). Neither days since last use nor CUD vs. healthy diagnosis were associated with any reliable feature after correction. Joint-history models indicated that the signal was better explained by longer-term use history than by recent use. Localization analyses indicated the connectivity effect was concentrated in dorsal thalamic regions. TMS-interaction and effective-connectivity follow-ups were null.

**Conclusions:** Reliability screening identified a thalamocortical control-network phenotype in CUD that tracks longer cocaine-use history rather than diagnosis or recent use. More broadly, this workflow offers a practical framework for screening candidate circuit-level psychiatric phenotypes for reliability.

## Introduction

Psychiatry still relies heavily on symptom-based diagnoses across many conditions, including substance use disorders such as cocaine use disorder (CUD). A long-term goal of the field is a more biologically informed psychiatry, in which disorders can be identified using measurable pathophysiology rather than symptom surveys alone. Considerable prior work has aimed to identify the neurobiological underpinnings of psychiatric disorders, including CUD, and has generated many candidate biological phenotypes.

However, for this work to become clinically useful, the field must move beyond generating new candidate patterns and instead determine which findings are reproducible. Many candidate brain signatures have been proposed, but the field still needs markers that are both biologically interpretable and demonstrably reliable. In cocaine use disorder (CUD), resting-state studies repeatedly implicate large-scale networks that support internally directed processing, salience allocation, and executive control (1-5). The next step is to identify which circuit-level findings hold up when they are tested with explicit reliability screens.

Thalamocortical circuitry may be especially well suited to yield clinically relevant circuit-level markers. Addiction models have long emphasized cortico-striatal-thalamo-cortical loops, yet human imaging studies have usually focused more on cortex and striatum than on the thalamus itself (6). This omission matters because the thalamus is a key organizer of large-scale cortical communication, not merely a relay structure. Functional-connectivity and graph-theory work suggests that it acts as an integrative hub across cortical systems (7). Modern anatomical and functional work also shows that the mediodorsal thalamus is internally heterogeneous and tightly linked to prefrontal functions, including executive control (8, 9). In cocaine dependence specifically, prior imaging has already shown evidence of abnormal thalamic connectivity patterns, but most of that work has treated the thalamus at a coarse scale or has used broad voxelwise or whole-brain approaches rather than a focused, hypothesis-driven thalamocortical panel (6, 10).

There is also a dynamic state reason to focus on this problem. Network studies in CUD have reported disrupted interactions among the default mode network (DMN), salience network, and executive-control network (2-4). A recent dynamic resting-state study went further and found that cocaine users showed a higher default-mode state occurrence rate and a higher probability of shifting from a salience state into a default-mode state. Both measures tracked dependence severity (11). In other words, prior work already suggests that CUD may involve not only altered average connectivity but also altered movement through recurring large-scale brain states.

The problem is that dynamic and connectivity features are not all equally trustworthy. Test-retest work in resting-state fMRI shows that many edge-level functional-connectivity measures are only modestly reliable, and dynamic summaries are often less reliable than static connectivity (12, 13). In practice, that means a study can generate many candidate features that look interesting but do not hold up when the same scan is split. This is especially relevant in CUD studies based on fMRI data, where motion, symptom heterogeneity, and limited sample sizes can make over-interpretation easy.

For this study, we focused on three cortical partners, the DMN core, control frontoparietal cortex (control FPC), and salience cortex, because prior CUD work repeatedly implicates these systems (1-4, 11). This study focused on their interaction with the thalamus due to its implication in prior addiction reviews, human thalamic connectivity work, and newer mediodorsal thalamus work that suggest that prefrontal-thalamic channels may be especially relevant to cognitive control and addiction-related behavior (6-10). Accordingly, we screened three types of candidate readouts: time spent in DMN-like and control-like states, transitions between those states, and thalamic connectivity with DMN core, control FPC, and salience cortex.

We also made two design choices to reduce circularity. First, the state model was trained only in healthy control participants (HC) and then reused unchanged in the disease analyses. Second, every candidate primary feature had to clear a split-half reliability gate before it entered primary inference. The main clinical targets were chosen as years since beginning cocaine use and days since last use. This allowed us to ask whether any of our hypothesized features act like a diagnosis marker, a recent-use marker, or a longer-term illness-history marker. More broadly, beyond the specific hypotheses tested here, this study provides a rigorous framework for distinguishing candidate brain phenotypes that remain robust under careful evaluation from those that fail to survive reliability screening.

## Methods and Materials

### Study overview and analytic samples

Resting-state Blood Oxygenation Level Dependent (BOLD) signal and T1-weighted anatomical images were obtained from two publicly available OpenNeuro datasets: SUDMEX-CONN and SUDMEX-TMS (14, 15). Raw BIDS-formatted data were downloaded to retrieve task-rest functional runs and their corresponding anatomical scans.

The datasets were minimally preprocessed with fMRIPrep 23.2.3. We then extracted 432 atlas-based regional time series (Schaefer-400 cortical parcels plus Tian S2 subcortical regions) and applied nuisance regression using motion parameters, global signal, high-pass basis functions, and aCompCor regressors, together with detrending and standardization (16, 17). No spatial smoothing or low-pass filtering were applied.

The raw CONN manifest contained 145 unique baseline participants and the paired TMS manifests contained 50 participants with both time-zero (T0) and time-one (T1) sessions available. After the workflow’s readiness and quality-control filters, the analytic discovery sample was 105 participants (46 HC, 59 CUD). The paired TMS extension sample included 44 participants with usable T0 and T1 data. The TMS sample was used only for follow-up analyses, and was not pooled with the discovery sample and did not contribute to state-model fitting.

### Clinical targets and covariates

The two primary clinical targets were years since beginning cocaine use (years.begin) and days since last use (days.last.use). Age at cocaine onset (coc.age.onset) was treated as a prespecified sensitivity target.

The shared nuisance covariates were age, sex, education, and mean framewise displacement (FD). Missing covariate values, when present, were mean-imputed column by column. Clinical target values were not imputed and were analyzed on an available-case basis. All covariates (age, sex, education, mean FD) and the imaging feature predictor were z-scored before entering the design matrix. Optional covariates were added only when their inclusion preserved a full-rank design matrix.

### Imaging input and thalamocortical circuit

Preprocessing yielded 432-node parcellated resting-state time-series files. The repetition time was 2.0 s. The thalamocortical circuit was defined programmatically from atlas label names.

The full circuit contained 13 named region-of-interest (ROI) summaries. Thalamic summaries were dorsal-anterior (DA), dorsal-posterior (DP), ventral-anterior (VA), ventral-posterior (VP), a dorsal composite (DA + DP), a ventral composite (VA + VP), and a whole-thalamus composite. The cortical and subcortical partner summaries were ventromedial prefrontal cortex (vmPFC), posterior cingulate cortex/precuneus (PCC/precuneus), their union (DMN core), control frontoparietal cortex (control FPC), salience cortex, and ventral striatum.

The HC-derived state model used a smaller 7-ROI subset: dorsal thalamus, ventral thalamus, vmPFC, PCC/precuneus, control FPC, salience, and ventral striatum. This subset was used only to define recurring states. Static and state-conditioned connectivity features were then computed from the broader thalamocortical circuit.

**Figure 1.**
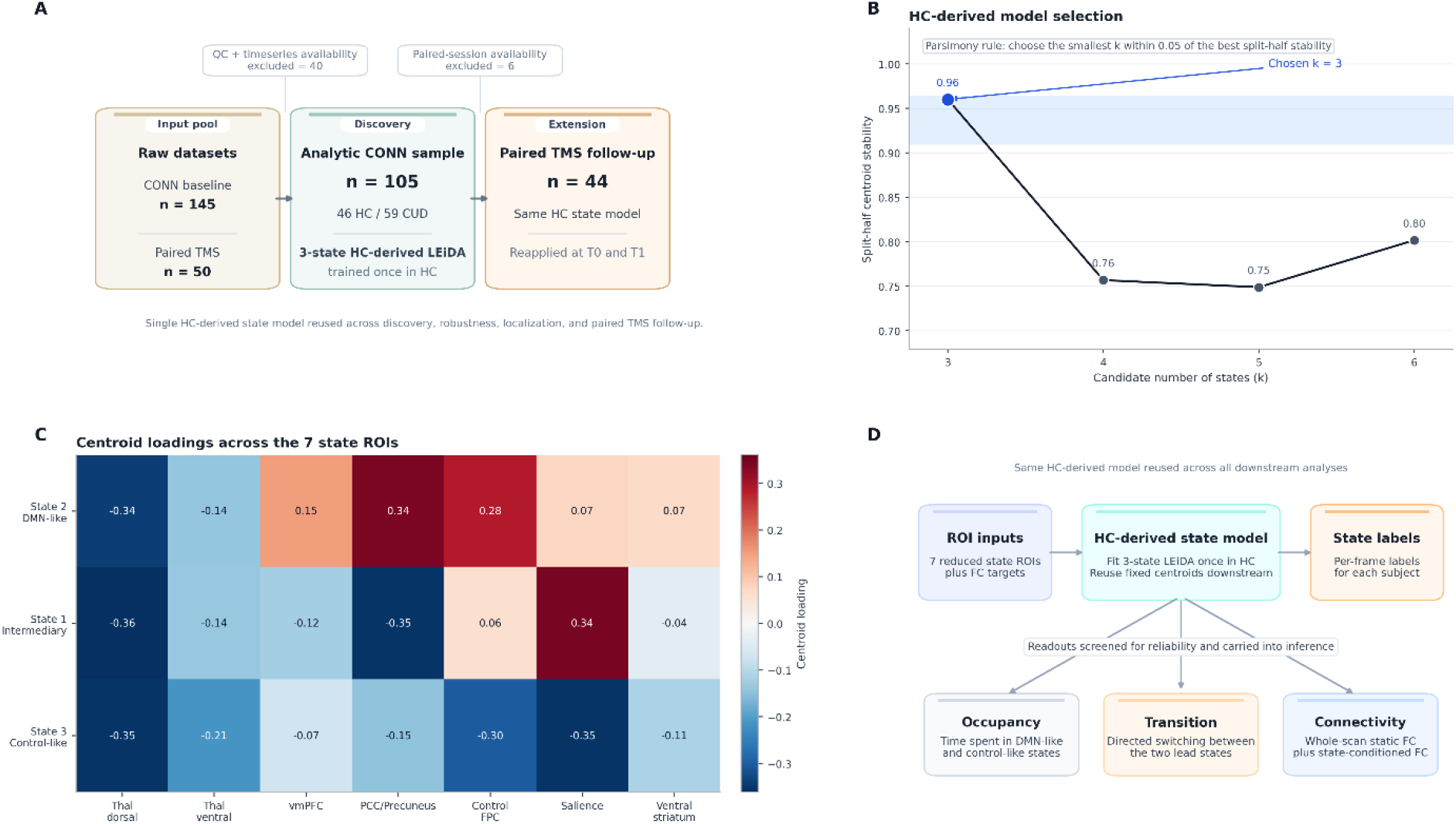
Study orientation and HC-derived thalamocortical state model. (A) Sample flow from raw datasets through quality control to the analytic CONN discovery sample (n = 105; 46 HC, 59 CUD) and paired TMS extension sample (n = 44). A single HC-derived LEiDA state model was trained once in healthy controls and reused across all downstream analyses. (B) Split-half centroid stability across candidate numbers of states (k = 3-6). A parsimony rule selected the smallest k within 0.05 of the best stability score, yielding k = 3 (stability = 0.96). (C) Centroid loading profiles across the 7 state ROIs for the 3 retained states: State 2 (DMN-like), State 1 (intermediary), and State 3 (control-like). (D) Workflow schematic. ROI inputs and per-frame state labels derived from the HC model were used to compute occupancy, transition, and connectivity readouts, each screened for split-half reliability before primary inference.

### HC-derived LEiDA state model

We fit the state model in HC only. For each HC participant, the 7 state-model ROI time series were standardized. At each time point, leading-eigenvector dynamics analysis (LEiDA) reduced the 7 × 7 phase-coherence matrix to one dominant spatial pattern, i.e. the leading eigenvector. That is to say, each frame was summarized by the main phase-locking pattern across the 7-ROI circuit.

Candidate models with k = 3, 4, 5, 6 states were compared by split-half centroid stability in HC. For each candidate k, HC participants were randomly split into two halves eight times. Frame-wise eigenvectors were pooled within each half, and k-means was fit separately in the two halves with 8 random starts and a maximum of 200 iterations. The resulting centroids were matched across all possible permutations to maximize their mean absolute correlation. The stability score for a given k was the average matched similarity across the eight repeats. We then selected the smallest k within 0.05 of the best stability score and refit the final k-means model in all HC state vectors.

State labels were assigned from the centroid loadings. The DMN-like state was the centroid whose absolute loadings favored vmPFC and PCC/precuneus over control FPC and salience. The control-like state was the converse. The remaining state was labeled intermediary.

### Prespecified feature families and reliability gate

Before any hypothesis testing, we generated a small prespecified primary feature family. It contained 11 person-level features:

1. DMN-like occupancy
2. Control-like occupancy
3. Transition probability from DMN-like to control-like
4. Transition probability from control-like to DMN-like
5. The difference between those two transition probabilities
6. Whole-thalamus static connectivity with DMN core
7. Whole-thalamus static connectivity with control FPC
8. Whole-thalamus static connectivity with salience
9. Whole-thalamus state-conditioned connectivity with DMN core during the DMN-like state
10. Whole-thalamus state-conditioned connectivity with control FPC during the control-like state
11. Whole-thalamus state-conditioned connectivity with salience during the control-like state

Occupancy was the fraction of frames assigned to a given state. Transition features came from the row-normalized transition matrix across consecutive frames. Static connectivity was the Pearson correlation between the relevant ROI-average time series across the full scan. State-conditioned connectivity used the same correlation, but only within frames assigned to the named state; these features were set to missing unless that state contained at least 12 frames. A separate localization family recomputed thalamus-control FPC static connectivity for DA, DP, VA, VP, dorsal composite, ventral composite, and whole-thalamus summaries.

We then applied a split-half reliability gate before any primary inference. Using the fixed HC-derived state model, the full feature engine was rerun on the first and second halves of each scan. Split-half agreement was summarized with Pearson r and Intraclass Correlation Coefficient (ICC(3,1)). A feature entered the primary analyses only if at least 8 participants had finite split-half values and ICC(3,1) was at least 0.40. Because half-scans are shorter, the minimum state count for split-half state-conditioned features was relaxed from 12 to 6 frames. The same reliability rule was later applied to the localization family and to the lag-1 effective-connectivity follow-up.

### Primary statistical models

The discovery stage used two primary model classes.

For group analyses, the imaging feature was the outcome and group (CUD = 1, HC = 0) was the predictor of interest. For dimensional analyses within CUD, the clinical target was the outcome and the imaging feature was the predictor of interest. In those within-CUD models, a positive beta therefore means that higher feature values were associated with larger target values.

In both model classes, age, sex, education, and mean FD were included when doing so preserved a full-rank design matrix. All models used ordinary least squares with HC3 robust standard errors. Significance of the coefficient of interest was tested with a Freedman-Lane permutation test using 1,000 permutations. Bootstrap 95% confidence intervals were computed from 500 resamples. The workflow also reported standardized coefficients.

Multiple testing was controlled with Benjamini-Hochberg false-discovery-rate (FDR) correction (18). For group models, correction was applied across modeled features. For within-CUD dimensional models, correction was applied separately within target across modeled features (years.begin or days.last.use).

### Follow-up analyses

We ran five follow-up analyses.

First, joint-history models asked whether the signal was better explained by longer-term use history or more recent use. We did this in two ways. In target-anchored models, a focal clinical target was predicted from a brain feature while adjusting for the other clinical target plus the usual nuisance covariates. In feature-anchored models, the brain feature itself was predicted from years.begin, days.last.use, and age, with sex, education, and mean FD added when rank permitted.

Second, localization analyses repeated the within-CUD dimensional model for thalamus-control FPC static connectivity computed separately for DA, DP, VA, VP, dorsal composite, ventral composite, and whole-thalamus summaries. FDR correction was applied within-target (years.begin or days.last.use) across partitions.

Third, motion-sensitivity analyses reran the primary within-CUD dimensional models after excluding participants with mean FD greater than 0.35 mm or 0.30 mm and after trimming the highest-motion 10% of participants.

Fourth, the TMS extension tested whether baseline features moderated treatment-related change in craving. For each baseline feature, we modeled T1 - T0 change in the Visual Analog Scale (VAS) or Cocaine Craving Questionnaire (CCQ) as a function of baseline feature, treatment, their interaction, baseline outcome, and nuisance covariates. We also computed an HC-normalization summary. Jointly reliable features were converted to HC-referenced z scores, collapsed to the mean absolute deviation from the HC reference at T0 and T1, and the change in that summary was compared between active and sham treatment while adjusting for baseline deviation.

Fifth, an exploratory lag-1 effective-connectivity follow-up fit a ridge-regularized VAR(1) model within each participant across 6 ROIs: dorsal thalamus, ventral thalamus, DMN core, control FPC, salience, and ventral striatum. Ridge regularization was fixed at alpha = 0.10. The requested lagged features were directional thalamus-control FPC terms, self-lag terms, and asymmetry terms.

## Results

### Sample flow and HC-derived state model

After quality control, the resting-state connectivity sample comprised 105 participants. The 46 healthy controls were on average 30.8 years old (SD = 7.7) with 12.9 years of education (SD = 3.5), and included 38 men and 8 women. The 59 participants with cocaine use disorder were on average 31.6 years old (SD = 7.1) with 11.0 years of education (SD = 3.0), and included 53 men and 6 women. The transcranial magnetic stimulation sample comprised 44 participants scanned at baseline and follow-up. The 20 participants receiving sham stimulation were on average 33.3 years old (SD = 8.7) with 12.9 years of education (SD = 2.7), and included 18 men and 2 women. The 24 participants receiving active stimulation were on average 35.8 years old (SD = 7.1) with 13.2 years of education (SD = 3.0), and included 20 men and 4 women.

Model selection clearly favored k = 3. Split-half centroid stability was 0.9601 for k = 3, 0.7568 for k = 4, 0.7489 for k = 5, and 0.8017 for k = 6. Under the prespecified parsimony rule, k = 3 was selected. Based on centroid loadings, the three resulting states were labeled DMN-like (State 2), intermediary (State 1), and control-like (State 3). The same HC-derived centroids were then reused in every downstream analysis.

### Reliability screen and primary findings

The reliability gate narrowed the primary feature set. Of the 11 prespecified primary features, only 5 passed the split-half ICC threshold: control-like occupancy, DMN-like occupancy, and whole-thalamus static connectivity with control FPC, DMN core, and salience. All transition features and all state-conditioned connectivity features failed the reliability gate and were excluded before inference.

**Figure 2.**
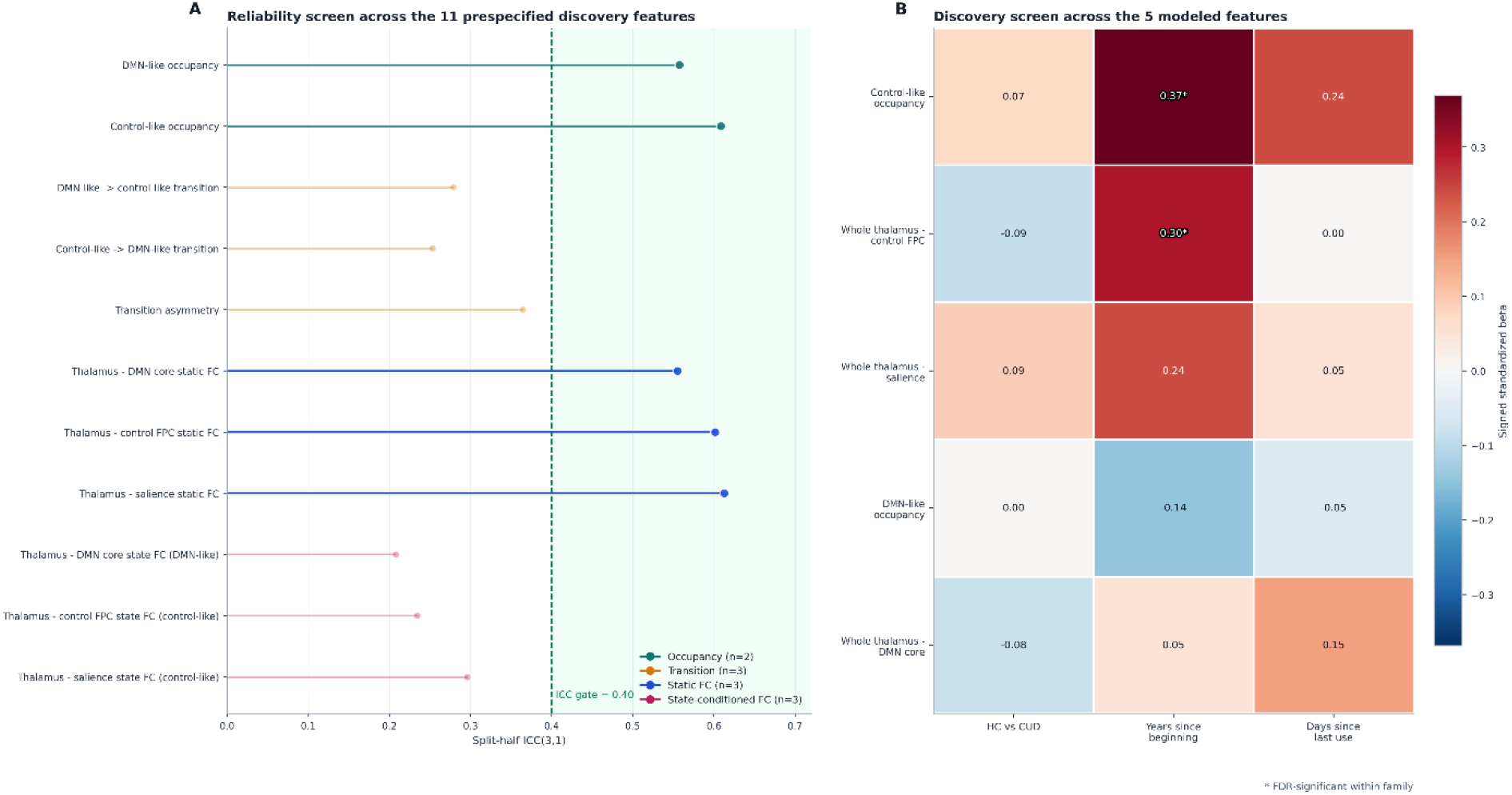
Reliability gate and discovery screen. (A) Split-half ICC(3,1) for all candidate gating features, ordered by reliability. The dashed line marks the ICC = 0.40 gate; features below this threshold were excluded from primary inference. (B) Discovery screen across the 5 modeled features. Cells show signed standardized coefficients for the HC-versus-CUD comparison and for the 2 primary clinical targets within CUD (years since beginning cocaine use and days since last use). Stars mark false-discovery-rate-significant associations. Only years since beginning cocaine use showed corrected findings, for control-like occupancy and whole-thalamus-control FPC static connectivity.

Within CUD, two reliable features tracked longer cocaine-use history. Participants with longer time since beginning cocaine use showed greater control-like occupancy (n = 58, beta = 2.4755, standardized beta = 0.3686, p_perm = 0.0010, q = 0.0050, 95% CI = 1.0409 to 3.7355) and stronger whole-thalamus-control FPC static connectivity (n = 58, beta = 1.9951, standardized beta = 0.2970, p_perm = 0.0070, q = 0.0175, 95% CI = 0.4660 to 3.1497).

No reliable feature was associated with days.last.use after FDR correction. The remaining three reliable features (DMN-like occupancy, whole-thalamus-DMN core static connectivity, and whole-thalamus-salience static connectivity) did not survive FDR correction for either primary target. None of the 5 reliable features differed between HC and CUD after FDR correction.

**Figure 3.**
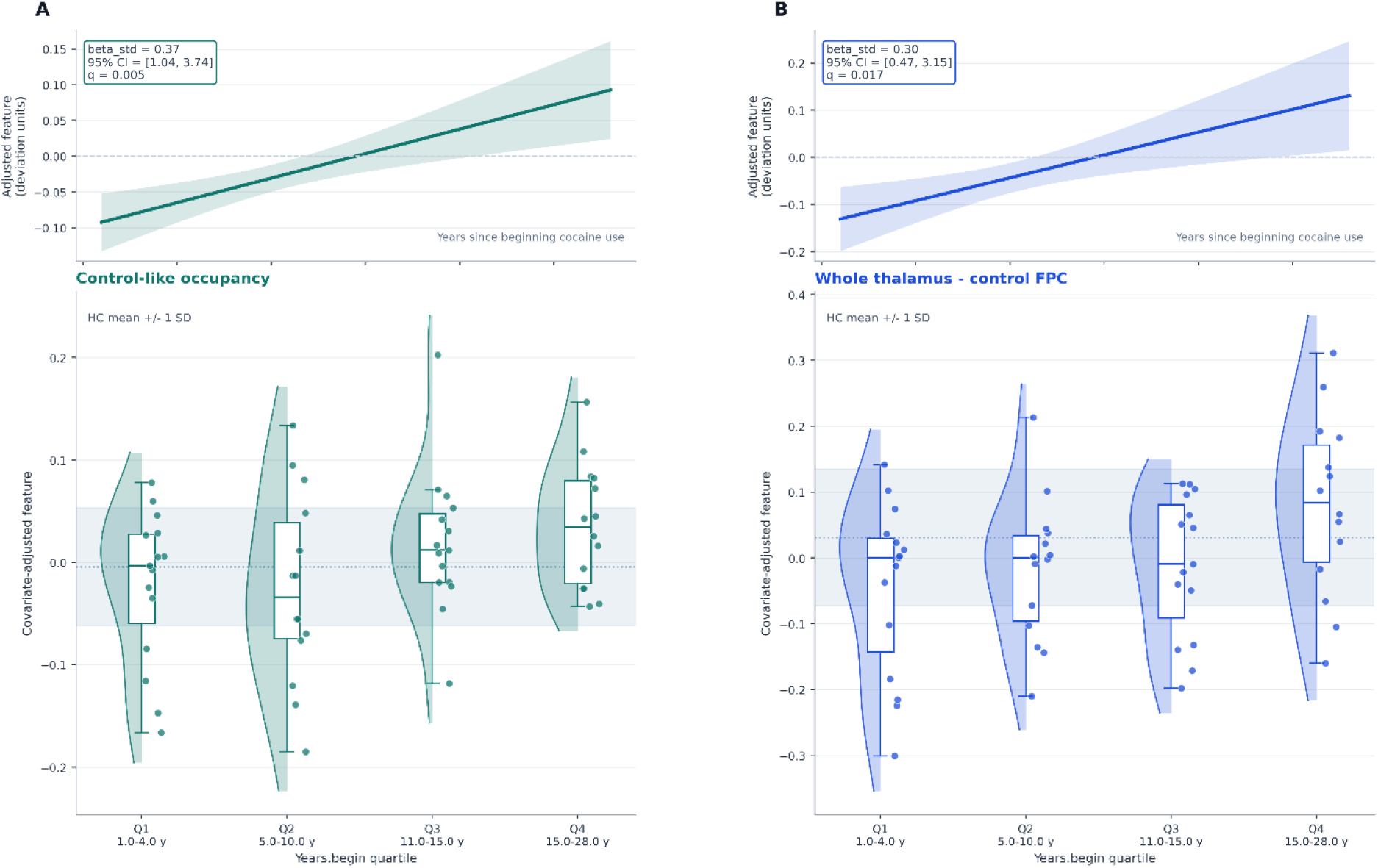
Dimensional phenotype visualization for the 2 lead years-since-beginning associations. Each column shows one false-discovery-rate-significant feature. Top row: covariate-adjusted marginal trend of the feature against years since beginning cocaine use within CUD, with a 95% confidence band. Bottom row: covariate-adjusted feature values split into quartiles of years since beginning cocaine use, displayed with half-violins, individual points, and boxplots. The shaded horizontal band marks the HC mean ± 1 SD for reference. (A) Control-like occupancy (standardized beta = 0.37, q = 0.005). (B) Whole-thalamus-control FPC static connectivity (standardized beta = 0.30, q = 0.017). Both features increase with longer cocaine-use history.

### Joint-history models

We next asked whether the years.begin signal simply reflected overlap with days.last.use. It did not.

In target-anchored models, control-like occupancy remained associated with years.begin after adjustment for days.last.use (n = 56, beta = 2.7894, standardized beta = 0.4170, p_perm = 0.0010, q = 0.0050, 95% CI = 1.3066 to 4.5182). The corresponding days.last.use model was null (q = 0.4146). The same pattern held for whole-thalamus-control FPC static connectivity: years.begin remained significant after adjustment for days.last.use (n = 56, beta = 2.1080, standardized beta = 0.3152, p_perm = 0.0090, q = 0.0225, 95% CI = 0.5004 to 3.1674), whereas days.last.use did not (q = 0.9580).

Feature-anchored models led to the same conclusion. Years.begin remained an independent predictor of control-like occupancy (n = 56, beta = 0.0486, standardized beta = 0.5744, p_perm = 0.0010, q = 0.0030, 95% CI = 0.0223 to 0.0714) and of whole-thalamus-control FPC static connectivity (n = 56, beta = 0.0693, standardized beta = 0.5218, p_perm = 0.0160, q = 0.0480, 95% CI = 0.0132 to 0.1093). Days.last.use did not explain additional variance in either lead feature.

### Motion robustness

Both lead years.begin effects were stable in every motion-sensitivity scenario.

For control-like occupancy, the association with years.begin remained positive and FDR significant in the FD <= 0.35 mm sample (n = 50, beta = 2.6552, q = 0.0150), the FD <= 0.30 mm sample (n = 45, beta = 2.5833, q = 0.0200), and the top-10%-trimmed sample (n = 50, beta = 2.6552, q = 0.0200).

For whole-thalamus-control FPC static connectivity, the same was true in all three scenarios: FD <= 0.35 mm (n = 50, beta = 2.2320, q = 0.0150), FD <= 0.30 mm (n = 45, beta = 2.2237, q = 0.0300), and top-10%-trimmed (n = 50, beta = 2.2320, q = 0.0150). In every scenario, the sign matched the full-sample estimate.

**Figure 4.**
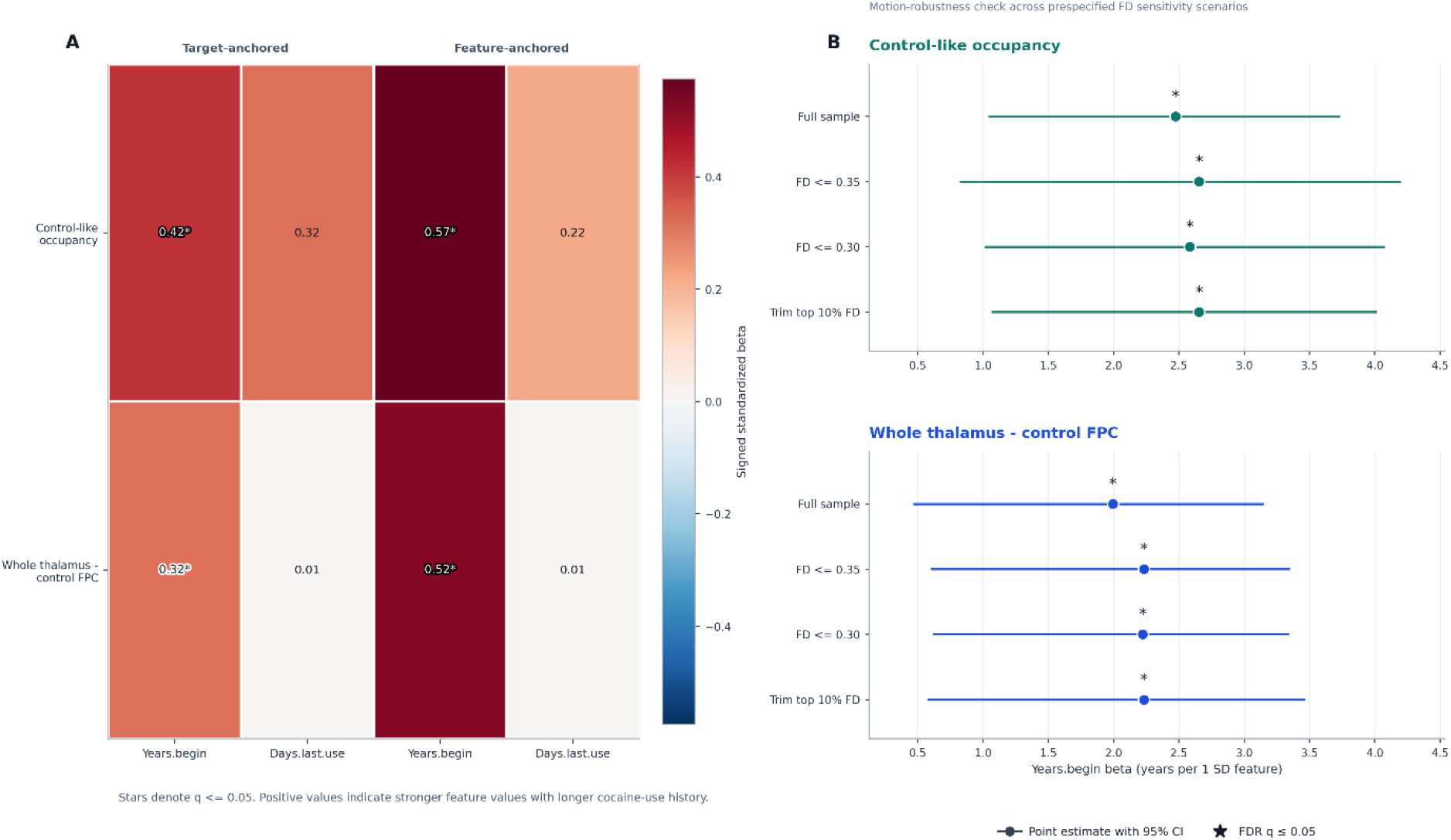
Joint-history adjudication and motion-robustness synthesis. (A) Joint-history coefficients for the 2 lead features under target-anchored models (left; each clinical target predicted from the brain feature while adjusting for the other target) and feature-anchored models (right; the brain feature predicted from both clinical targets simultaneously). Stars denote q <= 0.05. Years since beginning cocaine use remained informative in all lead models, whereas days since last use did not. (B) Motion-robustness forest plots for each lead feature. Points show the years-since-beginning coefficient in the full sample, the FD <= 0.35 mm sample, the FD <= 0.30 mm sample, and the top-10%-trimmed sample. Filled markers indicate the same sign as the full-sample estimate. Stars denote q <= 0.05. For both lead features, direction and significance were preserved across all scenarios.

### Thalamic localization

The thalamus-control FPC result was not evenly distributed across the thalamus. The strongest corrected associations with years.begin were concentrated in dorsal summaries.

FDR-significant localization effects were found for the dorsal composite (n = 58, beta = 2.1483, standardized beta = 0.3198, p_perm = 0.0050, q = 0.0315), the dorsal-posterior partition (n = 58, beta = 1.9886, standardized beta = 0.2961, p_perm = 0.0090, q = 0.0315), and the whole-thalamus summary (n = 58, beta = 1.9951, standardized beta = 0.2970, p_perm = 0.0140, q = 0.0326). Dorsal-anterior and ventral-posterior partitions were positive at the nominal level but did not survive correction (DA: beta = 1.5494, p_perm = 0.0350, q = 0.0531; VP: beta = 1.6088, p_perm = 0.0380, q = 0.0531). No thalamic partition showed an FDR-significant association with days.last.use.

**Figure 5.**
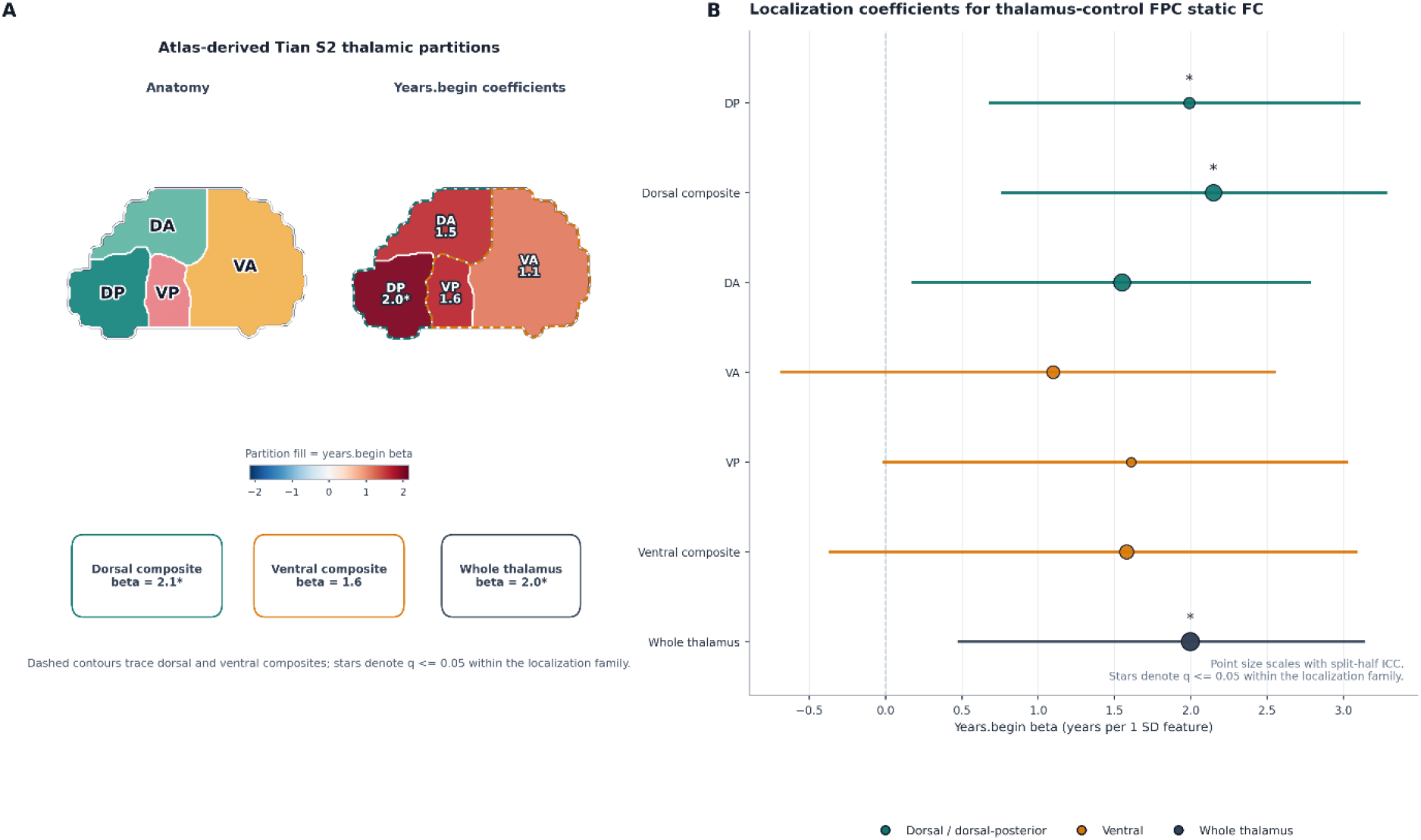
Dorsal thalamic localization of the years-since-beginning signal. (A) Left: atlas-derived Tian S2 thalamic partitions (DA, dorsal-anterior; DP, dorsal-posterior; VA, ventral-anterior; VP, ventral-posterior), with dashed contours indicating the dorsal and ventral composites. Right: the same partitions colored by the years-since-beginning coefficient for thalamus-control FPC static connectivity. (B) Forest plot of years-since-beginning coefficients for thalamus-control FPC static connectivity computed separately for each thalamic partition, the dorsal and ventral composites, and the whole-thalamus summary. Point size scales with split-half ICC. Stars denote q <= 0.05 within the localization family. Surviving effects were concentrated in dorsal summaries and in the whole-thalamus summary.

### Sensitivity target: age at cocaine onset

As a sensitivity analysis, earlier age at cocaine onset was associated with greater control-like occupancy (n = 58, beta = -2.3740, standardized beta = -0.4343, p_perm = 0.0080, q = 0.0400, 95% CI = -3.7356 to -1.0341). The corresponding whole-thalamus-control FPC association was negative but did not survive FDR correction. In the joint-history sensitivity models, age at onset did not remain independently informative once years.begin and days.last.use were included together.

### TMS extension and HC-normalization follow-up

The paired TMS extension did not support treatment moderation of the baseline phenotype. None of the 10 baseline-feature-by-treatment interaction models (5 features × 2 outcomes) survived FDR correction for either VAS or CCQ. The strongest nominal signal was baseline whole-thalamus-DMN core static connectivity for VAS change (beta = -1.2720, standardized beta = - 0.3550, p_perm = 0.0270), but this did not survive correction (q = 0.1349).

The HC-normalization summary was also null (hc_mean_abs_z: n = 44, beta = 0.0510, standardized beta = 0.0276, p_perm = 0.8322, q = 0.8322).

### Lag-1 effective-connectivity follow-up

None of the 10 requested lag-1 effective-connectivity features passed the reliability gate. The highest ICC(3,1) in that family was 0.2565 for the dorsal thalamus-control FPC asymmetry term, well below the prespecified threshold of 0.40. Because no requested lagged feature met the reliability criterion, no effective-connectivity association models were carried forward.

## Discussion

Psychiatry still lacks circuit-level markers that can sort patients by biology rather than by symptom lists. This study takes a step toward that goal in cocaine use disorder. Among eleven prespecified thalamocortical candidates, two features were sufficiently reliable and tracked longer cocaine-use history: time spent in a control-like state and static coupling between the thalamus and control frontoparietal cortex. The connectivity effect was concentrated in dorsal thalamic summaries. The study therefore identifies a measurable thalamocortical phenotype tied to clinically relevant variation within CUD.

The results emphasize the importance of prioritizing reliability and statistical stability when attempting to identify reliable biological phenotypes from neuroimaging data. In this dataset, the reliability gate sharply narrowed the field of plausible correlates. Transition features, state-conditioned connectivity features, and lag-1 effective-connectivity summaries were excluded before inference because they did not show adequate split-half stability. This filtering clarifies which classes of thalamocortical measures were stable enough to support interpretation in this dataset and which still require better upstream measurement or modeling.

The phenotype we found also fits with current models of addiction circuitry. Prior resting-state work in CUD has implicated executive-control, salience, default-mode, and frontostriatal systems (1-5, 11). The thalamus is positioned to coordinate information flow across these systems (6-10). Stronger dorsal thalamus-control FPC connectivity therefore points to altered organization in an associative thalamocortical channel that is relevant to cognitive control and long-term drug use.

Greater control-like occupancy should be interpreted cautiously. The label is derived from centroid loadings, so it identifies a recurring large-scale pattern rather than a direct mechanism. Several explanations remain plausible, including compensatory recruitment, a more rigid control-dominant configuration, or a premorbid trait related to vulnerability.

The follow-up analyses help place the phenotype on a clinical timescale. The main associations remained after adjustment for days since last use, which supports a longer-horizon signal linked to illness history. The screened features also did not separate HC from CUD after correction, and the paired TMS analysis did not show that baseline features moderated craving change. The present analysis therefore supports a phenotype of variation within CUD. Evidence for acute recency of use or treatment moderation was not observed in this dataset.

The study also has clear limitations. First, it is cross-sectional, so it cannot establish whether the phenotype reflects premorbid risk, cumulative neuroadaptation, compensation, or some mixture of the three. Second, years since beginning cocaine use is clinically meaningful, but it is still an indirect proxy for illness history. It does not measure dose, patterning, or total exposure. Third, the state labels are based on centroid loadings and should not be mistaken for direct physiology. Fourth, the discovery sample is moderate in size, and the paired TMS sample is smaller, which limits power for longitudinal or moderation analyses. Finally, the null effective-connectivity results may reflect low reliability of this simple lag-1 BOLD model rather than proof that directed interactions are absent.

Taken together, these analyses identify a thalamocortical control-network phenotype linked to cocaine-use history, with the connectivity component centered in dorsal thalamic partitions. More broadly, these analyses illustrate a replicable framework for evaluating candidate markers before they are interpreted as robust.

## Data Availability

This study used publicly available data from SUDMEX-CONN and SUDMEX-TMS. SUDMEX-CONN MRI data are available through OpenNeuro under accession ds003346, and SUDMEX-TMS MRI data are available through OpenNeuro under accession ds003037. Associated clinical and cognitive measures are available through the corresponding Zenodo records. The present code archive does not redistribute raw MRI data, fMRIPrep derivatives, or participant-level source time series. The analysis code, workflow documentation, and reproducibility outputs are archived on Zenodo at https://doi.org/10.5281/zenodo.19827264.

## Acknowledgements

This research was supported in part by grant GM-118039 from the Division of General Medical Sciences of the National Institutes of Health and a generous gift from the Ovarian Cancer Institute.

## Funding

This work was supported in part by grant GM-118039 from the Division of General Medical Sciences of the National Institutes of Health and by a generous gift from the Ovarian Cancer Institute. The funders had no role in study design, data analysis, interpretation, manuscript preparation, or the decision to submit the work for publication.

## Competing interests

The authors declare no competing interests.

## Ethics approval and consent to participate

This study was a secondary analysis of publicly available, de-identified data from the SUDMEX-CONN and SUDMEX-TMS datasets. For SUDMEX-CONN, the original study was conducted in accordance with the Declaration of Helsinki, was approved by the Ethics Committee of the Instituto Nacional de Psiquiatría “Ramón de la Fuente Muñiz,” and all participants provided verbal and written informed consent. For SUDMEX-TMS, the original study was approved by the Ethics Committee of the Instituto Nacional de Psiquiatría “Ramón de la Fuente Muñiz” under approval CEI/C/070/2016, was conducted in accordance with the Declaration of Helsinki, and was registered at ClinicalTrials.gov as NCT02986438. Participants provided informed consent in the original study.

## Consent for publication

The present manuscript does not include identifying images or individually identifying participant information.

## Data availability

This study used publicly available data from SUDMEX-CONN and SUDMEX-TMS. SUDMEX-CONN MRI data are available through OpenNeuro under accession ds003346, and SUDMEX-TMS MRI data are available through OpenNeuro under accession ds003037. Associated clinical and cognitive measures are available through the corresponding Zenodo records. The present code archive does not redistribute raw MRI data, fMRIPrep derivatives, or participant-level source time series.

## Code availability

The analysis code, workflow documentation, and reproducibility outputs are archived on Zenodo at https://doi.org/10.5281/zenodo.19827264.

## Author contributions

BE and JS conceived the study. BE designed and implemented the analytic workflow and performed the statistical analyses. BE and JS interpreted the results. BE drafted the manuscript. BE and JS reviewed and edited the manuscript, approved the final version, and agree to be accountable for the work.

## AI disclosure

The authors used artificial-intelligence tools (GPT-5.4 and 5.5, Claude Opus 4.6 and 4.7) to assist with code development, debugging, workflow organization, and manuscript editing for clarity. All analyses, code outputs, scientific interpretations, and manuscript text were reviewed and verified by the authors. The authors take full responsibility for the content of this work.

## Notes

### Competing Interest Statement

The authors have declared no competing interest.

### Author Declarations

This study was a secondary analysis of publicly available, de-identified data from the SUDMEX-CONN and SUDMEX-TMS datasets. For SUDMEX-CONN, the original study was conducted in accordance with the Declaration of Helsinki, was approved by the Ethics Committee of the Instituto Nacional de Psiquiatria Ramon de la Fuente Muniz, and all participants provided verbal and written informed consent. For SUDMEX-TMS, the original study was approved by the Ethics Committee of the Instituto Nacional de Psiquiatria Ramon de la Fuente Muniz under approval CEI/C/070/2016, was conducted in accordance with the Declaration of Helsinki, and was registered at ClinicalTrials.gov as NCT02986438. Participants provided informed consent in the original study.

